# Evidence of integrated health service delivery during COVID-19 in low- and middle-income countries: protocol for a scoping review

**DOI:** 10.1101/2020.07.23.20160721

**Authors:** Rachel Neill, Md Zabir Hasan, Priyanka Das, Vasuki Venugopal, Nishant Jain, Dinesh Arora, Shivam Gupta

## Abstract

**Introduction:** The importance of integrated, people-centered health systems has been recognized as a central component of achieving Universal Health Coverage. Integration has also been highlighted as a critical element for building resilient health systems that can stand the shock of health emergencies. However, there is dearth of research and systematic synthesis of evidence on the synergistic relationship between integrated health services and pandemic preparedness in low- and low-middle income countries (LMICs). Thus, the authors are organizing a scoping review aiming to explore application of integrated health service delivery approaches during the emerging COVID-19 pandemic in LMICs.

**Methods and analysis:** This scoping review adheres to the six steps for scoping reviews from Arksey and O’Malley (2005). Peer reviewed scientific literature will be systematically assembled utilizing a standardized and replicable search strategy from seven electronic databases, including PubMed, Embase, Scopus, Web of Science, CINAHL Plus, the World Health Organization’s Global Research Database on COVID-19, and LitCovid. Initially, the title and abstract of the collected literature, published in English from December 2019 to June 2020, will be screened for inclusion which will be followed by a full text review by two independent reviewers. Data will be charted using a data extraction form and reported in narrative format with accompanying data matrixes.

**Ethics and dissemination:** No ethical approval is required for the review. The study will be conducted from June to December 2020. Results from this study will provide a snapshot of the evidence currently being generated related to integrated health service delivery in response to the COVID-19 pandemic. The findings will be developed into reports and a peer-reviewed articles and will assist policy makers in making pragmatic and evidence-based decisions for current and future pandemic response.

**Article Summary:** *Strengths and limitations of this study:*

- The scoping review aims to uncover new evidence in response to the evolving pandemic and will not assess the quality of existing evidence
- The review will map a rapidly emerging evidence base in response to COVID-19 from seven different electronic databases that can be used to inform response and recovery strategies in LMICs
- The early research and published evidence on the COVID-19 pandemic was focused in high-income and upper-middle income countries, thus, we expect evidence of from LMICs may be scarce
- The scoping review is limited to peer review publications that were originally written in English or have translated versions

## INTRODUCTION

Coronavirus disease 2019 (COVID-19) was first reported on 31 December 2019 in Wuhan, the capital city of Hubei province in China. Sprouting from a local outbreak, COVID-19 emerged as one of the most significant pandemics of the last century [1,2]. Within six months, over 10 million confirmed COVID-19 cases had been detected worldwide with over 500 thousand deaths [3]. The COVID-19 pandemic has taken a toll on the health systems of emerging economies of South-east Asia, with India, Pakistan, and Bangladesh in the top twenty countries for total cases [3].

This unprecedented pandemic has put an enormous amount of financial, administrative, and logistical stress on the health sector [4,5], including the health systems of low and low-middle-income countries (LMIC). Within the midst of this systemwide shock, preparedness, response, and recovery endeavors need to be adopted by the health system to control the COVID-19 effectively [6]. These adaptations include expanding surveillance systems to identify potential cases and contact trace, the development of communication strategies between health systems and populations to share of credible and accurate information, and sustaining routine health care services across settings and levels of the health system. However, due to the often segmented and vertically organized health system in LMICs, these adaptations often come with heavy costs in time, financial, human resources, and de-prioritization of routine healthcare services. [7]

### Integrated health service delivery system

The importance of integrated, people-centered health systems was globally affirmed in 2016 with an adopted resolution of the sixty-ninth World Health Assembly [8] and is increasingly recognized as a central component of achieving Universal Health Coverage [9]. This research adopts the definition of integrated health service delivery (IHSD) system from the World Health Organization (WHO) Regional Office for Europe:

“An approach to strengthen people-centered health systems through the promotion of the comprehensive delivery of quality services across the life-course, designed according to the multidimensional needs of the population and the individual and delivered by a coordinated multidisciplinary team of providers working across settings and levels of care… … with feedback loops to continuously improve performance and to tackle upstream causes of ill health and to promote well-being through intersectoral and multisectoral actions.” [10]

The above definition represents a comprehensive view of IHSD and captures integration from the multiple perspectives of health systems stakeholders (patients, providers, managers, and policymakers). Within the broad definition, four dimensions of integration are explored: organizational, functional, service, and clinical [11]. These dimensions are summarized in Table 1, adapted from Lewis et al. 2008 [11].

**Table 1.** Dimensions of Integration.

| **Dimension of Integration** | **Description of the Dimensions** |
| --- | --- |
| Organizational | Integration of organizational units, whether formally through a merger/acquisition, informally through collaboration and referral, or financially by a single purchaser or payor. Organizational integration can be vertical (across levels of care) or horizontal (within levels of care) |
| Functional | Integration of clinical and non-clinical functions |
| Service | Integration of different services within a single care team or organizational unit |
| Clinical | Integration of standardized clinical processes, guidelines, and/or protocols used within/across providers |

Source: Lewis RQ, Rosen R, Goodwin N, et al. *Where next for integrated care organisations in the English NHS?* London: The Nuffield Trust 2010.
https://www.nuffieldtrust.org.uk/files/2017-01/where-next-integrated-care-english-nhs-web-final.pdf

### Integrated health service delivery for pandemic preparedness, response, and recovery

The World Health Organization’s 2017 guide on influenza risk management outlines four pandemic phases [6] – (i) interpandemic, (ii) alert, (iii) pandemic, and (iv) transition, that align with three components of risk assessment – (a) preparedness, (b) response, and (c) recovery. Literature specific to pandemic preparedness also highlights aspects of system integration, albeit from a broader perspective than the definition of IHSD used here.

It is possible that typologies of integration recommended to prepare and respond to a health emergency could relate to, or even advance, broader movements towards an IHSD system. For example, a coordinated approach – including comprehensive risk management, multidisciplinary health sector collaboration, and building community resilience – is recommended for emergency management [6]. This requires planning to surge capacity, developing triage systems, managing the continuity of essential health services, and prevention of secondary effects are emphasized for pandemic preparedness [6]. It is highly likely that integration across the components of health systems, levels of care, and across types of providers will prove necessary during the COVID-19 pandemic.

The interdependency between the IHSD system and pandemic preparedness is especially pertinent in LMICs. The surge of infection related to Severe Acute Respiratory Syndrome Coronavirus 2 (SARS-CoV-2) spared the LMICs in the early stages of the pandemic. However, as COVID-19 incidence is increasing rapidly through developing countries like India, the pandemic has drawn attention towards the required integration of a range of health services and supply chains that are often under-resourced in LMICs, such as ambulatory care, mental health services, and oxygen provision. Historically health systems in the LMICs were structured and sustained as vertical and disease-focused, and we acknowledge that significant success were achieved. However, the same fragmentation in the health system could create challenges to the coordinated pandemic preparedness, response, and recovery effort against COVID-19.

Bolstered by the 40th anniversary of the Alma Alta Declaration and the movement towards Universal Health Coverage, many LMIC health systems have moved away from a vertical disease focus towards IHSD approaches [12,13]. This shift has also been increasingly advocated by bilateral and multilateral donor agencies and international organizations as a means to respond to the changing burden of disease (from infectious to non-communicable diseases), improve people-centeredness, and increase efficiency [12–14]. This review can identify any emerging evidence on where integrated care has improved health systems outcomes during COVID-19 in LMICs and provide guidance on the further testing of emerging approaches.

## RATIONALE OF THE REVIEW

The importance of integration (OR harmonization OR coordination) – both within the health delivery system and between public health and health services delivery systems – has been highlighted as key to building resilient health systems that can respond to health emergencies [15–17]. Each of these aspects could potentially relate to, or be strengthened by, IHSD systems, but this explicit linkage appears under-addressed by existing literature. This study aims to explore the evidence and application of integrated health service delivery across the pandemic phases and risk assessment components in LMICs using a scoping review methodology.

Scientific research related to COVID-19 is emerging as rapidly as the pace of the pandemic itself [18,19] (Figure 1). We can assume that the evidence for the IHSD system during the COVID-19 pandemic is rapidly evolving as the epidemiologic profile and disease burden changes worldwide. Moreover, there is an acute lack of exploration of the published evidence focusing on the application of integrated health service delivery in LMICs. Thus, a scoping review is an appropriate design to map existing literature, summarize it, and identify gaps [20]. Scoping studies are also appropriate to contextualize existing knowledge and gaps within a policy and practice context [21].

Exploring IHSD within the context of the COVID-19 pandemic provides a unique opportunity to examine whether LMICs are utilizing integrated approaches in response to an external shock to their health system, what approach(es) are being used, and what recommendations are emerging. The review will also identify any emerging evidence on where integrated care has improved health systems outcomes during COVID-19 in LMIC settings and provide guidance on the adoption of various typologies. This scoping review will enable a mapping of the evidence from published literature as it relates to the IHSD system. Results from this study will be disseminated to support policy makers and practitioners in LMICs to support pragmatic and evidence-based decisions for current and future pandemic response.

## OBJECTIVES

Aligning with the aim of the overarching aim of this study, this scoping review has three objectives:

1. Investigating the characteristics of the IHSD system commonly appearing in the COVID-19 literature generated from LMICs
2. Exploring the operational approaches of IHSD being utilized during COVID-19 preparedness, response, and recovery within the health systems of LMICs
3. Identifying the emerging recommendations on the IHSD system for pandemic preparedness, response, and recovery during the COVID-19 pandemic from LMICs

## METHODS AND ANALYSIS

The methodology for this scoping review follows Arksey and O’Malley’s six stages for scoping reviews [20] and adheres to the checklist of Preferred Reporting Items for Systematic Reviews and Meta-Analyses’ Extension for Scoping Reviews (PRISMA-ScR) [22,23]. An overview of each stage of the review follows.

### Stage one: conceptualizing research question

The overarching research question leading this scoping review is: *How are the health systems of LMICs utilizing IHSD approaches to prepare for and/or respond to the COVID-19 pandemic?* This research question aligns itself with the research objective mentioned in the above section.

### Stage two: identification of relevant literature

Followed by the development of the research objective, a comprehensive and replicable literature search strategy is being structured to extract the references of the relevant peer reviewed articles from literature repositories (Table 2). To implement this process, first, the research team has identified key literature from PubMed and Google Scholar to select keyword and index terms and develop the search terms. Next, using the keywords and search terms, the study will conduct a comprehensive search across seven electronic databases.

**Table 2.** Database and their website address which was included in the scoping review.

| No | Database name | Website link |
| --- | --- | --- |
| 1 | PubMed | [ncbi.nlm.nih.gov/pubmed/](https://pubmed.ncbi.nlm.nih.gov/) |
| 2 | Embase | [embase.com](https://www.embase.com/) |
| 3 | Scopus | [scopus.org](https://www.scopus.org/) |
| 4 | Web of Science | [webofknowledge.com](https://www.webofknowledge.com/) |
| 5 | CINAHL Plus | https://health.ebsco.com/products/cinahl-plus |
| 6 | LitCovid | https://www.ncbi.nlm.nih.gov/research/coronavirus/ |
| 7 | WHO COVID-19 literature database | https://search.bvsalud.org/global-literature-on-novel-coronavirus-2019-ncov/ |

The search strategy of the electronic database consists of four concepts: (i) Integrated care, (ii) pandemic preparedness, (iii) COVID-19, and (iv) names of the countries belong to the low- and low-middle-income group according to the World Bank Classification [24]. Using these concepts, the initial search strategy developed for PubMed generated only 15 records published between 01 December 2019 to 22 April 2020 (Search conducted on 22 April 2020). The proposed search strategy is presented in Table 3 and detailed in the supplementary material of this protocol.

**Table 3:**
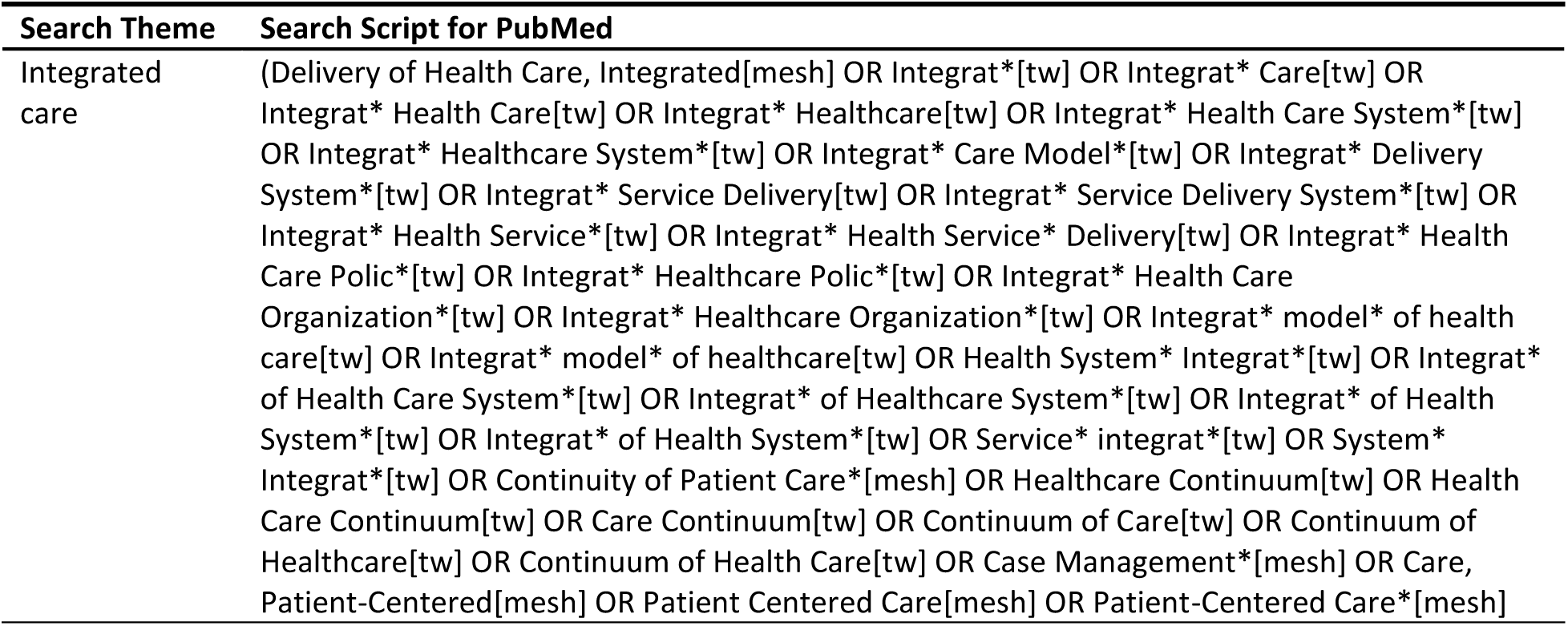
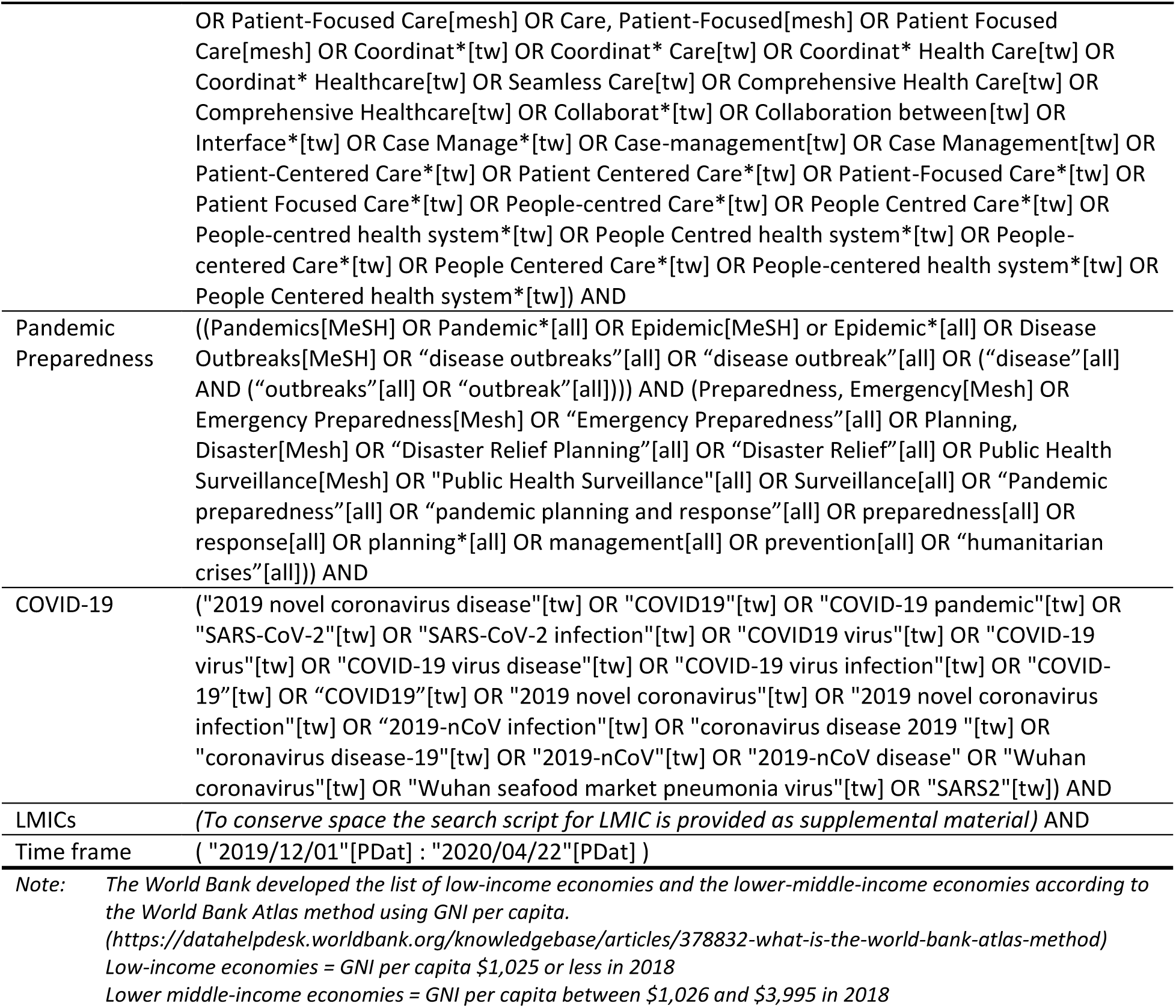
PubMed literature search script for the scoping review (implemented on 22 April 2020)

As the literature on COVID-19 is rapidly changing, a brief title and abstract scan was conducted to review the performance of the search strategy. Upon this brief review, the research team concluded that few articles were published from LMICs until 22 April 2020. Thus, a decision was taken to conduct a second round of literature search on 12 June 2020. The second implementation of the search strategy in PubMed generated 92 records, published between 01 December 2019 to 12 June 2020 (Search conducted on 12 June 2020).

After implementing the search strategy in all seven databases, the title and abstracts will be downloaded, and citations will be imported into Covidence systematic review software (covidence.org). At this stage, we will remove the duplicates and organize the search records to review their titles and abstracts.

### Stage three: study selection

During the third stage, all input articles will be screened utilizing the pre-determined inclusion and exclusion criteria found in Table 3. In line with Arksey and O’Malley’s methodological framework for scoping reviews, the inclusion and exclusion criteria represent a broad view of the subject, and the evidence characteristics may be satisfied by a range of study designs and methodologies [20].

Two independent reviewers will screen the title and abstract of each imported document against the Table 4 criteria, and any conflicting recommendations will be reviewed and adjudicated by a third reviewer for consistency. Next, the full text of the initially selected articles will be screened using the same criteria by two independent reviewers, followed by review and adjudication of conflicting recommendations by the third reviewer. As scoping reviews are often iterative [20,25], any suggested modifications to the inclusion or exclusion criteria will be reviewed by the entire research team with the senior member making final decisions regarding necessary modifications. If changes to the criteria are agreed upon, all previously excluded documents will be re-screened to ensure appropriate inclusion or exclusion against the modified criteria.

**Table 4.**
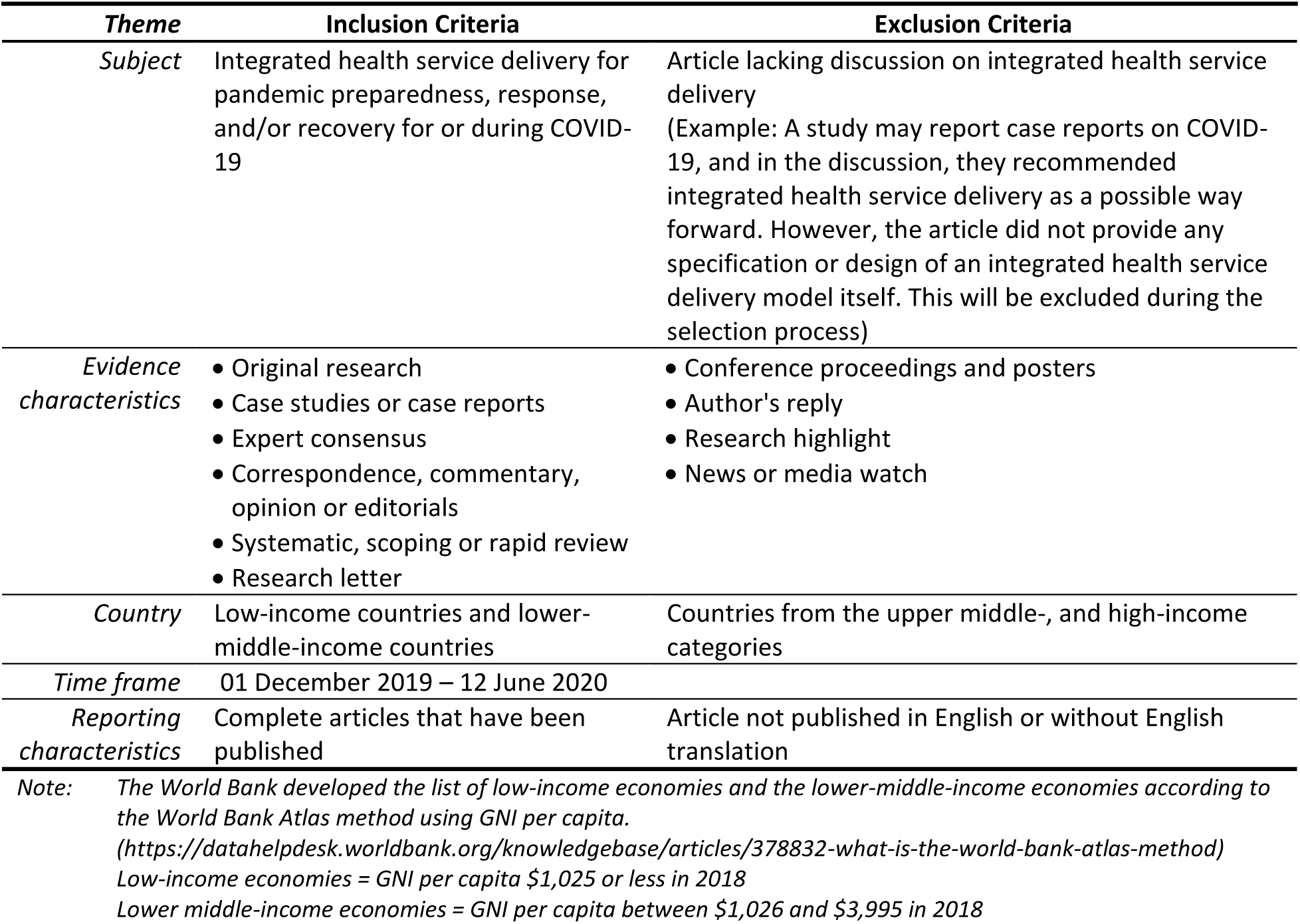
Inclusion and exclusion criteria for the study selection process of the scoping review.

### Stage four: charting data

Eligible articles from the full-text review will be re-examined, and the relevant data from the articles will be charted using a data extraction form. Table 5 provides an overview of the initial overview of data elements, which will be extracted for the study (see the supplementary materials for the full extraction form and overview of terms).

**Table 5.** Data extraction template.

| **Data extraction themes** | **Data elements that will be extracted from each eligible article** |  |
| --- | --- | --- |
| *Study characteristics* | • Database<br>• Title<br>• Authors<br>• Year<br>• Type of article<br>• Country name(s) or global focus | • Country type (World Bank classification)<br>• WHO region<br>• Study populations<br>• Study location<br>• Study design and methodology<br>• Framework utilized (if any) |
| *Dimensions of Integrated Care and Pandemic Preparedness* | • Definition of IHSD<br>• Pandemic phase when IHSD implemented<br>• IHSD related risk assessment (preparedness, response, and recovery)<br>• Typologies of integration<br>• Type(s) of service(s) integrated (if applicable) | • Integration mechanism (if applicable)<br>• Integration structure (if applicable)<br>• Integration intensity (if applicable)<br>• Incentives for integration (if applicable)<br>• Timing of integration (if applicable)<br>• Organizational and operational components of integration |
| *Intersection with COVID-19* | • Provided details on COVID-19 pandemic specific to a country (if applicable)<br>• Facilitators of integration<br>• Barriers to integration<br>• Positive effects of integration | • Negative consequences of integration<br>• Recommendations – COVID-19 specific<br>• Recommendations – health system (non-COVID-19 specific) |

*Note:* The World Bank developed the list of low-income economies and the lower-middle-income economies according to the World Bank Atlas method using GNI per capita. https://datahelpdesk.worldbank.org/knowledgebase/articles/378832-what-is-the-world-bank-atlas-method Low-income economies = GNI per capita \$1,025 or less in 2018 Lower middle-income economies = GNI per capita between \$1,026 and \$3,995 in 2018

### Stage five: reporting the results

Charted information will be analyzed thematically and reported in a narrative format using tables and figures. Per standardized methodology for scoping reviews, the quality of evidence will not be assessed during the reporting process [20]. Data displays and matrixes will be utilized to explore aspects of integration vis a vis the risk assessment phases of pandemic response. Dedoose (dedoose.com) will be utilized to explore clusters of findings geographically and across phases of pandemic response.

### Stage six: expert consultation

Expert consultation is an optional stage proposed by Arksey and O’Malley [20]. We believe that there is an inherent value of the expert consultation to translate the findings of this scoping review. Though we intend to perform a brief expert consultation, implementation of this stage will be based on the feasibility and progression of the scoping review.

The potential roster of experts can be developed from the Department of International Health of Johns Hopkins Bloomberg School of Public Health (JHSPH), Johns Hopkins Center for Health Security (JHCHS) and Deutsche Gesellschaft für Internationale Zusammenarbeit (GIZ) Gmbh.

## TIMELINE

Stages one and two of this review was initiated from April 2020 and iterated until June 2020. Stage three – title and abstract screening and full-text review – will be conducted from July to September 2020. Stages four through six are expected to be completed by November 2020. The estimated completion timeline of the scoping review is December 2020.

## PATIENT AND PUBLIC INVOLVEMENT

No patients or members of the public were involved or consulted in the development of the study or its intended execution.

## ETHICS AND DISSEMINATION

Ethical review is not required for the scoping review. Only publicly available secondary data will be utilized for the review, and no primary data will be collected. A report and a peer-reviewed publication will be developed for broader dissemination synthesized evidence of the scoping review. There may be additional opportunities to disseminate the findings at conferences or webinars to support the COVID-19 response in low and low-middle income countries.

## Data Availability

This study is developed from publicly available secondary data. The scoping review is registered on OSF.io with the Registration DOI 10.17605/OSF.IO/KY9PX (osf.io/yk7gu).

## AUTHOR CONTRIBUTIONS

RVN, MZH, and SG developed the review’s objectives; MZH developed the search strategy and conducted the search; RVN developed the manuscript under the supervision of SG and with contribution from MZH, PD, VV, NJ, and DA. All authors contributed to manuscript revision and read and approved the protocol for publication.

## ACKNOWLEDGMENT

We would like to thank Welch Medical Library of Johns Hopkins University, specifically collaborating Informationist Donna Hesson, for providing support to conduct the scoping review and for assistance in developing the search terms.

## ETHICAL APPROVAL

No ethical approval is required for the study.

## REFERENCES

1 Ravelo JL, Jerving S. COVID-19 — a timeline of the coronavirus outbreak. Devex. 2020.https://www.devex.com/news/sponsored/covid-19-a-timeline-of-the-coronavirus-outbreak-96396 (accessed 26 Jun 2020).

2 World Health Organization. Rolling updates on coronavirus disease (COVID-19). World Health Organization. 2020.https://www.who.int/emergencies/diseases/novel-coronavirus-2019/events-as-they-happen (accessed 26 Jun 2020).

3 Johns Hopkins University. Coronavirus COVID-19 Global Cases. Johns Hopkins Coronavirus Resource Center. 2020.https://coronavirus.jhu.edu/map.html (accessed 2 Apr 2020).

4 Ehrlich H, McKenney M, Elkbuli A. Protecting our healthcare workers during the COVID-19 pandemic. Am J Emerg Med Published Online First: 17 April 2020. doi:10.1016/j.ajem.2020.04.024

5 Farrell TW, Francis L, Brown T, et al. Rationing Limited Healthcare Resources in the COVID-19 Era and Beyond: Ethical Considerations Regarding Older Adults. J Am Geriatr Soc 2020;68:1143–9. doi:10.1111/jgs.16539

6 World Health Organization. Pandemic Influenza Risk Management: A WHO guide to inform and harmonize national and international pandemic preparedness and response. Geneva: 2017. https://apps.who.int/iris/bitstream/handle/10665/259893/WHO-WHE-IHM-GIP-2017.1-eng.pdf;jsessionid=FF0E44DE342CCEF9F0A31E1EFB14C8E8?sequence=1 (accessed 26 Jun 2020).

7 Mills A. Health Care Systems in Low- and Middle-Income Countries. New England Journal of Medicine 2014;370:552–7. doi:10.1056/NEJMra1110897

8 World Health Organization Secretariat. Framework on integrated, people-centred health services. 2016.https://apps.who.int/gb/ebwha/pdf_files/WHA69/A69_39-en.pdf?ua=1 (accessed 26 Jun 2020).

9 World Health Organization. Framework on integrated people-centred health services. WHO. http://www.who.int/servicedeliverysafety/areas/people-centred-care/framework/en/ (accessed 26 Jun 2020).

10 World Health Organization: Regional Office for Europe. Integrated care models: an overview. Copenhagen, Denmark:: WHO Regional Office for Europe 2016. http://www.euro.who.int/ data/assets/pdf_file/0005/322475/Integrated-care-models-overview.pdf (accessed 5 Jan 2020).

11 Lewis RQ, Rosen R, Goodwin N, et al. Where next for integrated care organisations in the English NHS? London:: The Nuffield Trust 2010. https://www.nuffieldtrust.org.uk/files/2017-01/where-next-integrated-care-english-nhs-web-final.pdf

12 Chaitkin M, Blanchet N, Su Y, et al. Integrating Vertical Programs into Primary Health Care: A Decision-Making Approach for Policymakers. Washington, DC: : Results for Development 2019. https://www.r4d.org/wp-content/uploads/Integrating-Vertical-Programs-into-PHC-synthesis-report-FINAL.pdf (accessed 26 Jun 2020).

13 Mounier-Jack S, Mayhew SH, Mays N. Integrated care: learning between high-income, and low- and middle-income country health systems. Health Policy Plan 2017;32:iv6–12. doi:10.1093/heapol/czx039

14 Lê G, Morgan R, Bestall J, et al. Can service integration work for universal health coverage? Evidence from around the globe. Health Policy 2016;120:406–19. doi:10.1016/j.healthpol.2016.02.007

15 Commission on a Global Health Risk Framework for the Future, Secretariat National Academy of Medicine. Strengthening Public Health as the Foundation of the Health System and First Line of Defense. National Academies Press (US) 2016. https://www.ncbi.nlm.nih.gov/books/NBK368392/ (accessed 26 Jun 2020).

16 Nuzzo JB, Meyer D, Snyder M, et al. What makes health systems resilient against infectious disease outbreaks and natural hazards? Results from a scoping review. BMC Public Health 2019;19:1310. doi:10.1186/s12889-019-7707-z

17 Kruk ME, Myers M, Varpilah ST, et al. What is a resilient health system? Lessons from Ebola. The Lancet 2015;385:1910–2. doi:10.1016/S0140-6736(15)60755-3

18 Chen Q, Allot A, Lu Z. Keep up with the latest coronavirus research. Nature 2020;579:193–193. doi:10.1038/d41586-020-00694-1

19 World Health Organization. Global research on coronavirus disease (COVID-19). World Health Organization. 2020.https://www.who.int/emergencies/diseases/novel-coronavirus-2019/global-research-on-novel-coronavirus-2019-ncov (accessed 26 Jun 2020).

20 Arksey H, O’Malley L. Scoping studies: towards a methodological framework. International Journal of Social Research Methodology 2005;8:19–32. doi:10.1080/1364557032000119616

21 Anderson S, Allen P, Peckham S, et al. Asking the right questions: scoping studies in the commissioning of research on the organisation and delivery of health services. Health Res Policy Syst 2008;6:7. doi:10.1186/1478-4505-6-7

22 Tricco AC, Lillie E, Zarin W, et al. PRISMA Extension for Scoping Reviews (PRISMA-ScR): Checklist and Explanation. Annals of Internal Medicine 2018;169:467–73. doi:10.7326/M18-0850

23 Munn Z, Aromataris E, editors. Joanna Briggs Institute Reviewer’s Manual. The Joanna Briggs Institute 2017. https://reviewersmanual.joannabriggs.org/

24 World Bank Group. World Bank Country and Lending Groups. The World Bank. 2020.https://datahelpdesk.worldbank.org/knowledgebase/articles/906519-world-bank-country-and-lending-groups (accessed 26 Jun 2020).

25 Micah DJ Peters, Christina Godfrey, Patricia McInerney, et al. Chapter 11: Scoping reviews. In: Aromataris E, Munn Z, eds. Joanna Briggs Institute Reviewer’s Manual,. JBI 2020. https://reviewersmanual.joannabriggs.org/

